# Genome-wide association study of Idiopathic Pulmonary Fibrosis susceptibility using clinically-curated European-ancestry datasets

**DOI:** 10.1101/2025.01.30.25321017

**Authors:** Daniel Chin, Tamara Hernandez-Beeftink, Lauren Donoghue, Beatriz Guillen-Guio, Olivia C Leavy, Ayodeji Adegunsoye, Helen L Booth, CleanUP-IPF Investigators of the Pulmonary Trials Cooperative, William A Fahy, Tasha E Fingerlin, Bibek Gooptu, Ian P Hall, Simon P Hart, Mike R Hill, Nik Hirani, Simon R Johnson, Naftali Kaminski, Jose Miguel Lorenzo-Salazar, Shwu-Fan Ma, Robin J McAnulty, Mark I McCarthy, Amy D Stockwell, Toby M Maher, Ann B Millar, Philip L Molyneaux, Maria Molina-Molina, Vidya Navaratnam, Margaret Neighbors, Justin M Oldham, Helen Parfrey, Gauri Saini, Ian Sayers, X Rebecca Sheng, Iain D Stewart, Mary E Strek, Martin D Tobin, Moira KB Whyte, Maria C Zarcone, Yingze Zhang, Fernando Martinez, Brian L Yaspan, Carl J Reynolds, David A Schwartz, Carlos Flores, Imre Noth, R Gisli Jenkins, Richard J Allen, Louise V Wain

## Abstract

**Background:** Idiopathic pulmonary fibrosis (IPF) is a rare, incurable lung disease with a median survival of 3-5 years after diagnosis. Treatment options are limited. Genetic association studies can identify new genes involved in disease that might represent potential new drug targets, and it has been shown that drug targets with support from genetic studies are more likely to be successful in clinical development. Previous genome-wide association studies (GWAS) of IPF susceptibility have identified more than 20 signals implicating genes involved in multiple mechanisms, including telomere dysfunction, cell-cell adhesion, host defence immunity, various signalling pathways and, more recently, mitotic spindle assembly complex.

**Aim:** To leverage new datasets and genotype imputation to discover further genes involved in development of IPF that could yield new pathobiological avenues for exploration and to guide future drug target discovery.

**Methods:** We conducted a GWAS of IPF susceptibility including seven IPF case-control studies comprising 5,159 IPF cases and 27,459 controls of European ancestry, where IPF diagnosis was made by a respiratory clinician according to international guidelines. Genotypes were obtained from Whole Genome Sequencing (WGS) or from array-based imputation to the TOPMed WGS reference panel. New signals were replicated in independent biobanks with IPF defined using Electronic Healthcare Records. Bayesian fine-mapping was performed to identify the most likely causal variant(s) and bioinformatic investigation undertaken to map associated variants to putative causal genes.

**Results:** We identified three novel genetic signals of association with IPF susceptibility. Genes prioritised by functional evidence at these signals included *MUC1*, which encodes a large transmembrane glycoprotein and known biomarker of lung fibrosis, and *NTN4* encoding Netrin-4 whose known roles include angiogenesis. The third signal may map to *SLC6A6*, a taurine and beta-alanine transporter gene, previously implicated in retinal, cardiac and kidney dysfunction.

**Conclusion:** Our study has identified new associations not previously identified by previous large biobank-based studies thereby highlighting the value of utilising clinically-curated IPF case-control studies, and new genotype imputation. We present new evidence for disease-driving roles of *MUC1* and of endothelial cell and vascular changes in IPF.

## INTRODUCTION

Idiopathic pulmonary fibrosis (IPF) is a chronic, progressive lung disease thought to result from an aberrant response to lung injury, culminating in an exaggerated healing response with excessive deposition of extracellular matrix in the interstitium (1) (2). This incurable lung disease affects more than 7 in 100,000 people with poor survival (median survival of 3-5 years after diagnosis) (3) and has limited treatment options (4). IPF is incompletely understood and is significantly influenced by both genetic and environmental factors, with a high heritability and polygenic aetiology (5) (6).

Genome-wide association studies (GWAS) assess genetic variants from across the genome for their association with disease. Previous GWAS have highlighted more than 20 independent genetic signals linked to IPF susceptibility, implicating pathways such as telomere dysfunction, cell-cell adhesion, host defence, TGF-β signalling, and mitotic spindle assembly (5) (7) (8) (9) (10) (11). Whole genome sequencing (WGS) technologies provide comprehensive genomic coverage, but remain cost prohibitive for large studies. Instead, reference panels derived from large WGS datasets allow for improved imputation of unmeasured variants, including those with low allele frequencies, providing a cost-effective solution for increased genome-wide coverage (12).

Although each genetic signal has a small individual effect, except for the gain-of-function *MUC5B* promoter variant (6) (13), targeting the pathways conferred by genetic associations could offer significant therapeutic potential, as drug targets supported by genetic evidence studies have a higher likelihood of success in clinical development (14). This is driving efforts to increase the discovery of IPF-associated genes and to refine existing signals to identify the most likely causal variants for molecular and mechanistic evaluation.

For this study, we re-imputed previously published studies using a more recent imputation panel that enables measurement of three times as many variants as our previous studies and aggregated new datasets comprising clinically-curated IPF cases, and controls, to improve the quality and coverage of genotyping and to increase sample size for new gene discovery while offering more precise risk estimates. Our findings implicate new loci for IPF susceptibility.

## METHODS

### Datasets

Seven independent case–control studies were analysed: Colorado (15), US (9), UK (10), IPF Job Exposures Study (IPF-JES) (16), Genentech (17), United States, United Kingdom, and Spain (UUS) (11) and Study of Clinical Efficacy of Antimicrobial Therapy Strategy Using Pragmatic Design in Idiopathic Pulmonary Fibrosis-University of California, Davis (CleanUP-UCD) (18) datasets (**Figure 1**). All studies included unrelated European-ancestry individuals, with cases of IPF diagnosed according to the American Thoracic Society and European Respiratory Society guidelines (19) (20) (21). Written informed consent and ethics approval were properly obtained for all studies, following the World Medical Association’s Code of Ethics (Declaration of Helsinki) and approved by the appropriate institutional review board or Research Ethics Committee. More information on each study can be found in the supplementary material.

**Figure 1.**
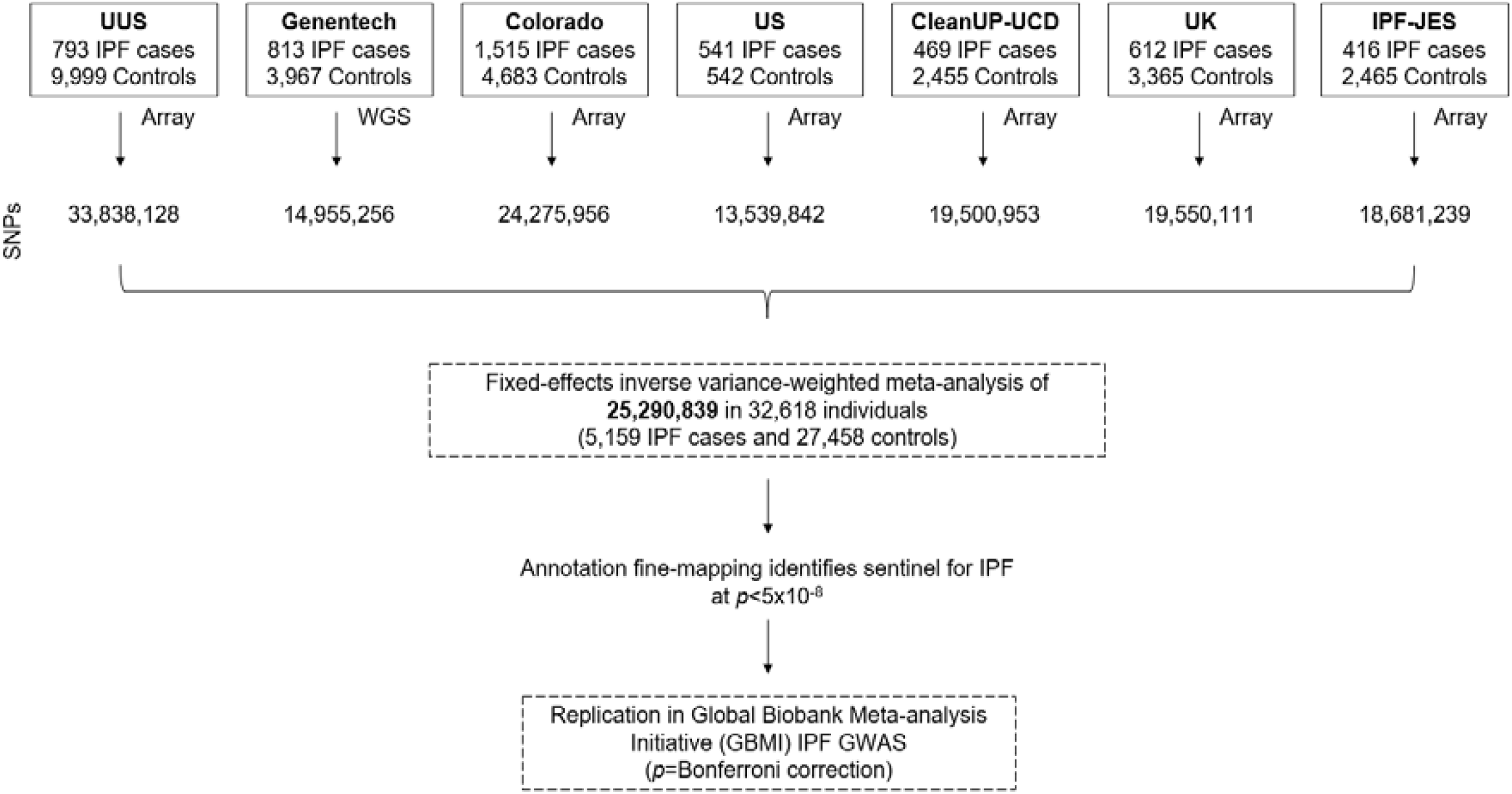
Study overview. SNPs: Single Nucleotide Polymorphisms. WGS: Whole-genome sequencing data.

### Genotyping and quality controls

The Colorado, IPF-JES, UK, US, UUS, and CleanUP-UCD studies had been genotyped using SNP arrays (supplementary material). Quality control measures included filtering for low call rates, sex mismatches, heterozygosity, non-European genetic ancestry, relatedness, and ensuring no overlap between studies. These six studies were imputed using the TOPMed WGS reference panel (GRCh38) via the TOPMed Imputation Server (12). For the Genentech study, genotypes were obtained through WGS on the Illumina HiSeq X Ten platform with an average read depth of 30X. Related individuals and those with call rates below 10% were excluded from the analysis. Additional details are available in previously published studies (15) (9) (10) (16) (17) (11) (18) and the supplementary material.

### Association analysis and meta-analysis

For six studies (Colorado, IPF-JES, UK, US, UUS, and CleanUP-UCD), genome-wide association analyses were conducted using logistic regression with PLINK v2 (22) adjusting each study for the first ten principal components to correct for population stratification. Variants with a poor imputation quality (r2<0.5) and minor allele count (MAC) ≤3 were removed. Association analysis for the Genentech study was performed using logistic regression with PLINK v1.9 (22) including sex, age, and five genetic-ancestry principal components as covariates.

The seven datasets were meta-analysed using inverse-variance weighted fixed effects meta-analysis with METAL (23), excluding variants that did not pass quality control in at least two studies. No minor allele frequency filter was applied. We estimated the genomic inflation factor using LDSC (24), and we applied a correction when this factor exceeded 1.10 in the meta-analysis. Data for chromosome X were available for four studies (UUS, Colorado, CleanUP-UCD, and UK) with GWAS and meta-analysis performed as described above (see supplementary material). For chromosome X, we applied a correction when classic lambda factor exceeded 1.15 in the meta-analysis.

### Signal selection

Independent association signals were selected after performing conditional analysis with COJO-GCTA v1.90.2 (25), and based on a *p* value threshold of <5.0×10^−8^. Forest plots were prepared using the forestplot R v4.1.3 package and visually reviewed for outlying study signals. Signals were excluded if the significance (*p* value) of the association in any individual study was lower than the meta-analysis significance (i.e., if combining data across studies served to reduce the association rather than support it). We sought replication of new signals using an independent subset of the Global Biobank Meta-analysis Initiative (GBMI) IPF GWAS (26) comprising 6,257 cases defined using Electronic Healthcare Records (EHR), and 947,616 controls, all of European ancestry. We defined a replication threshold of *p*<0.05 with Bonferroni correction for the number of new signals tested. We annotated the sentinel variants with ANNOVAR v07.06.20 (27) and Ensembl Variant Effect Predictor (VEP) v.105, and we identified potentially causal variants using the Wakefield Bayes factor method to calculate the posterior inclusion probability (PIP) of each variant to define a 95% credible set (28). We performed look-ups for all SNPs previously reported for association with IPF at genome-wide significance (*p*<5×10^−8^).

### *In silico* functional assessment

We performed a comprehensive functional impact assessment using empirical data from several integrated software tools and datasets for the sentinel SNPs and for each variant in the credible set with a PIP>0.1. We interrogated gene expression Quantitative Trait Locus (eQTLs) from the Genotype-Tissue Expression Project (GTEx) v8 (29) to identify whether IPF variants were associated with expression of any genes in lung, cultured fibroblasts or whole blood (see supplementary material for further details). We used coloc (30) R v4.1.3 package to investigate whether the same causal variant was driving both the genetic association with IPF risk and an association with gene expression changes and report shared signals with a colocalisation probability hypothesis 4 (H4) >0.7. We functionally annotated the highlighted regions based on active chromatin marks (i.e., DHS, eRNAs, ATACseq, CHIPseq, H3K27ac) (31) and searched for mouse orthologs of the human genes where knockout resulted in a measurable respiratory phenotype (https://www.mousephenotype.org/). We used Open Targets Genetics to identify additional supporting data for variant to gene mapping and for phenome-wide association study (PheWAS) analysis.

## RESULTS

We analysed 5,159 IPF cases and 27,459 controls of European ancestry, and a total of 25,290,839 autosomal variants (**Figure 2 & Table S1**). For X chromosome, association analyses were performed for 23,890 individuals (3,388 IPF cases and 20,502 controls), and 261,736 variants. There was no evidence of inflated test statistics (**Figure 2 & Figure S1**). After conditional analyses, we identified 37 autosomal independent significant signals (*p*<5×10^−8^), three of which were supported by multiple studies and had not been previously reported (**Table 1 & Figure 3**). All three signals were replicated in the independent GBMI IPF GWAS (*p*<0.0125 (**Table 1**). Only one signal (3p25.1) included a single SNP that accounted for more than 50% of the posterior probability of being causal (rs112271207 PIP=0.691) (**Table S2**). All signals overlapped open chromatin marks/regions (ATACseq or DHS) with evidence of enhancer activity (CHIPseq: H3K27ac) (**Table 2 & Table S3**). No signals reached the genome-wide significance threshold for the X chromosome analysis.

**Table 1.**
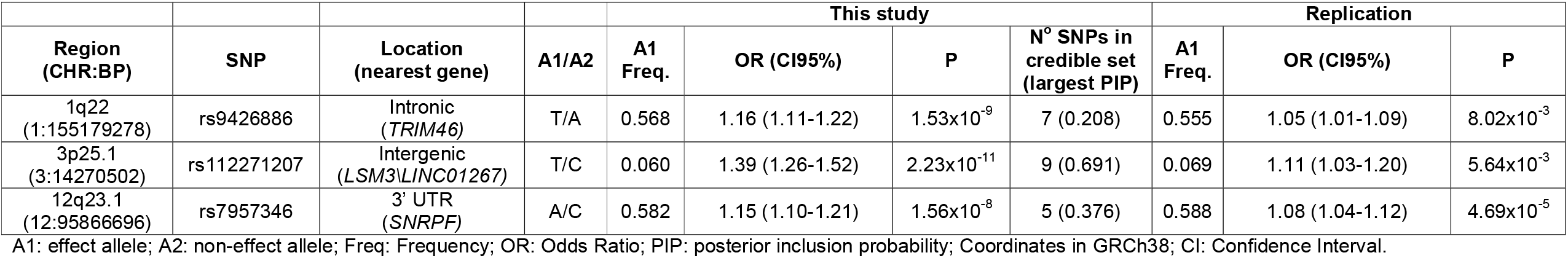
New genome-wide significant association signals.

**Table 2.** Summary of variant to gene mapping and functional information.

| Signal* | Location<br>(nearest gene) | Gene implicated by eQTL<br>(GTEx: tissue, highest<br>posterior probability of<br>shared causal variant) | Other gene evidence<sup>#</sup> | PheWAS (aligned to<br>IPF risk allele)<sup>#</sup> | Open Chromatin** |
| --- | --- | --- | --- | --- | --- |
| 1q22<br>(rs9426886) | Intronic<br>(*TRIM46*) | *MUC1* (whole blood,<br>0.616) | *THBS3*, *ADAM15* (sQTL<sup>1</sup>;<br>DHS-promoter correlation<sup>2</sup>) | increased haematocrit,<br>decreased urate | DHS; H3K27ac |
| 3p25.1<br>(rs112271207) | Intergenic<br>(*LSM3**LINC01267*) | NA | *SLC6A6* (PCHiC<sup>3</sup>) | decreased lung function | ATACseq; H3K27ac |
| 12q23.1<br>(rs7957346) | 3' UTR<br>(*SNRPF*) | *NTN4* (lung, 0.649) | *NTN4* (sQTL<sup>2</sup>) | decreased lung function | ATACseq; H3K27ac |

<sup>#</sup>Open Targets Genetics: 1. Splice QTL (source: GTEx); 2. DHS-promoter capture (source: Thurman et al 2012 (57)); 3. Promotor Capture HiC in blood-derived endothelial precursors, macrophages, megakaryocytes, naive B cells and neutrophils (from Open Targets Genetics, source: Javierre et al 2016 (58)).
\*\*Biddie et al 2023 (31). \*Minimum study-level imputation quality Rsq for each variant: rs9426886:0.936, rs112271207:0.929, rs7957346:0.991.
See Table S3 for full results.

**Figure 2.**
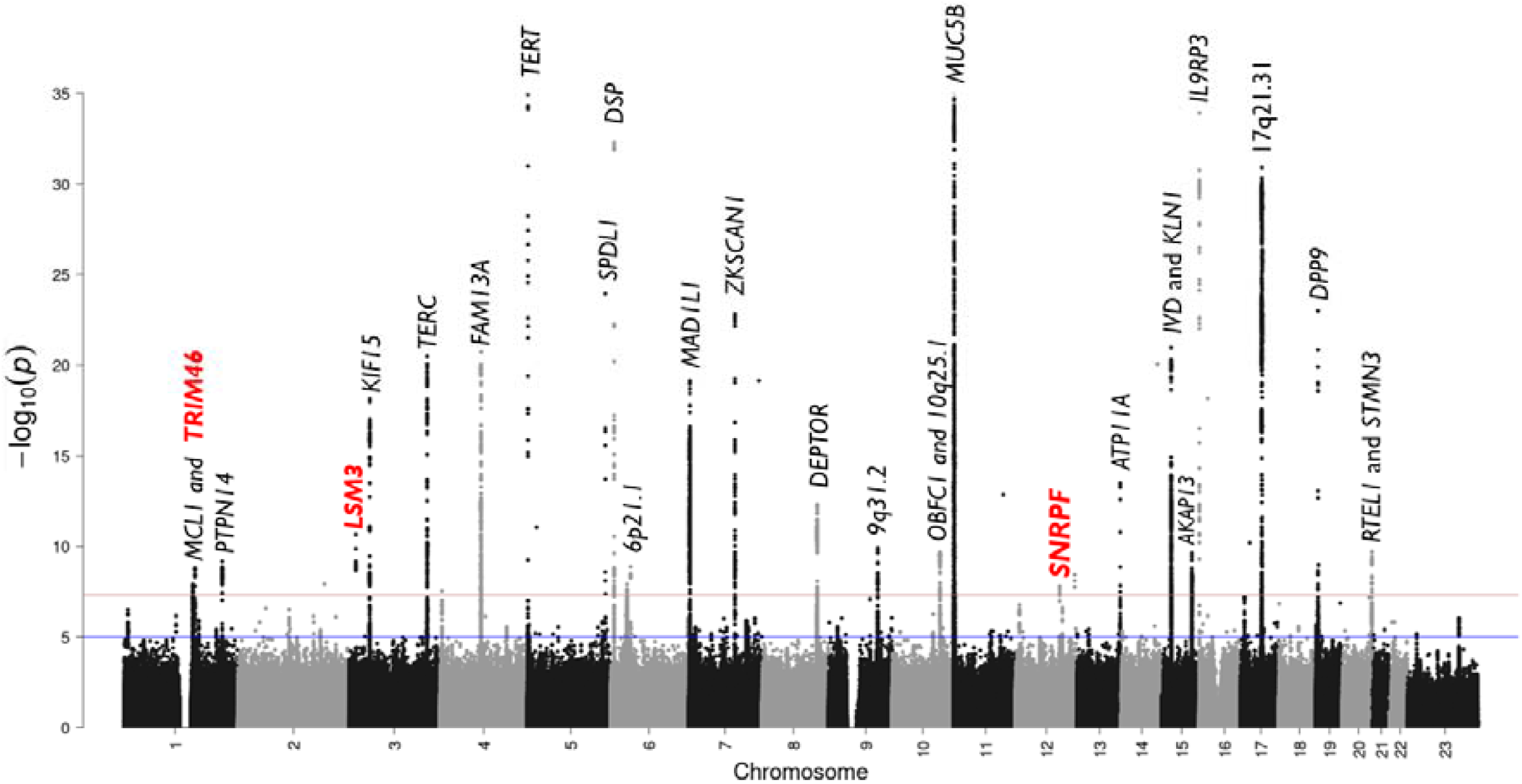
Manhattan plot of meta-analysis results. The y-axis shows the transformed p-values (–log10[p-value]) while the x-axis represents chromosome positions. The horizontal red line corresponds to the genome-wide threshold (*p*=5.0×10^−8^) and the blue line shows the suggestive significance threshold (*p*=5.0×10^−5^). The genomic inflation factor of the meta-analysis results did not show major deviations from the null hypothesis of no association. Newly discovered nearest annotated genes are highlighted in red and bold. The plot has been truncated at *p*=1.0×10^−35^. The most significant previously reported *MUC5B* signal (rs35705950) has a p<5.0×10^−121^.

**Figure 3.**
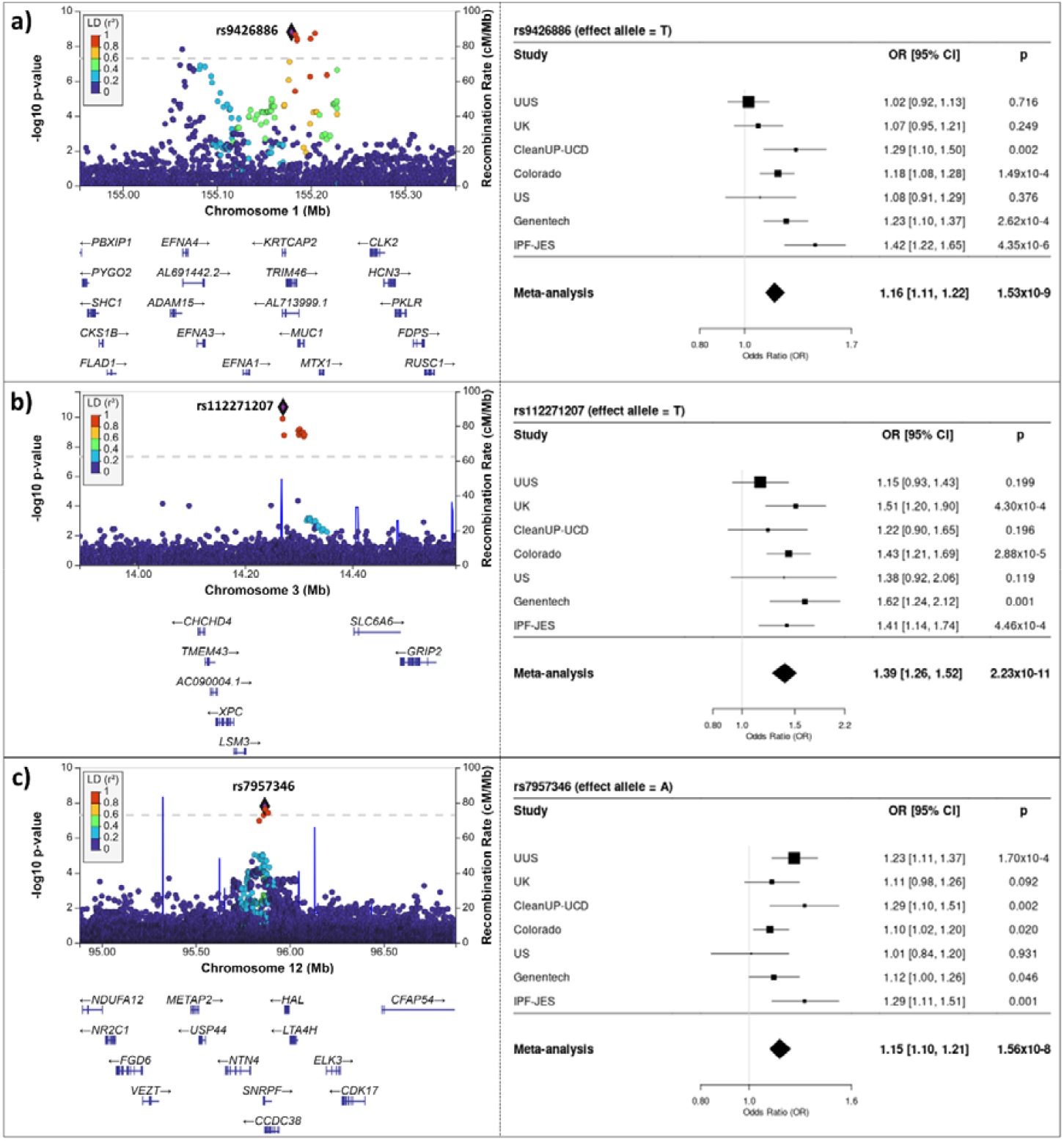
Region plots and forest plots of the three signals of interest: a) 1q22 region; b) 3p25.1 region; and c) 12q23.1 region. In region plots, the y-axis shows the transformed p-values (-log10[p-value]), while the x-axis represents chromosomal positions (GRCh38). The genome-wide significance threshold (*p* =5.0×10^−8^) is indicated by the horizontal dashed line. Linkage disequilibrium values (r2) are presented according to the LD colour scheme of the upper left legend. Plots were generated using LocusZoom (http://locuszoom.org/).

There were 32 previously reported IPF susceptibility signals and 31 were represented by variants in our dataset (**Table S4)** (an intronic signal, rs539683219, in the *PSKH1* gene could not be assessed and had previously been shown to be unique to East Asian ancestry populations (7)). All but three signals (in or near *DNAJB4, FSTL5* and *TPI1P2*) met a Bonferroni-corrected threshold of *p*<1.56×10^−3^ in our study and all but seven signals reached genome-wide significance. These seven included five previously reported association signals (in or near *DNAJB4, FSTL5, TPI1P2, GPR157*, and *FKBP5*) whose initial discovery had been enabled by a multi-ancestry study design) (7) and two signals (at 10q25.1 and *STMN3*) previously reported in a meta-analysis of a subset of the studies included here (5).

The most statistically significant new signal (rs112271207, 3p25.1) was a common (MAF 6%) intergenic variant located 91.6 kb downstream of *LSM3* and 77.6 kb upstream of *LINC01267*. This SNP had previously been associated at near-genome-wide significance with lung function (*p*=6.1×10^−8^) with the allele associated with increased risk of IPF (T) associated with decreased lung function. The variant was not associated with gene expression in GTEx. The Open Targets Genetics V2G annotation prioritised a member of a family of sodium and chloride ion dependent transporters (*SLC6A6)* (132 kb downstream) as the most likely causal gene based on chromatin interaction data (**Table S3**).

The second most significant signal (rs9426886, 1q22) was located within an intron of the *TRIM46* gene, which is involved in the formation of parallel microtubule bundles. Although the sentinel variant was significantly associated with expression of several genes in cultured fibroblast, lung and whole blood (**Table S5**), the highest colocalisation probability was 0.616 for *MUC1* in whole blood. *MUC1* encodes a cell-surface glycoprotein. The IPF risk allele (T) associated with increased expression of *MUC1* in whole blood. This signal was also associated with urate levels (*p*=1.5×10^−101^) and haematocrit measurement (*p*=4.7×10^−41^) with the allele associated with increased IPF risk associated with increased haematocrit measurement, and decreased urate levels.

Finally, the signal at 12q23.1 (rs7957346) had the highest probability of colocalisation for expression of Netrin 4 (*NTN4*) in lung tissue (posterior probability = 0.649) and had previously been associated with lung function with the IPF risk increasing allele (A) associated with decreased lung function (*p*=7.8×10^−45^). *NTN4* encodes Netrin 4; netrins are a family of proteins involved in cell and axon migration during development and link to angiogenesis and vascularization.

## DISCUSSION

Using our large clinically-curated IPF case-control resources, together with new imputation to the TOPMed Reference panel, we report novel associations that extend our previous genetic findings for IPF susceptibility and implicate new genes involved in its pathobiology.

The signal at 1q22 suggestively implicated *MUC1* which encodes a cell surface glycoprotein well known to be important in several lung diseases and infections (32). The MUC1 glycoprotein comprises three domains; an extracellular domain, a transmembrane domain, and a cytoplasmic tail. The extracellular domain becomes cleaved during alveolar epithelial damage and is commonly known as Krebs Von Den Lungen-6 (KL-6). KL-6 has been widely recognised as a potential prognostic biomarker of lung fibrosis as it is elevated in the serum and bronchioalveolar lavage fluid of patients with IPF. Studies have shown that KL-6 may also drive fibrosis through promotion of fibroblast activation and their differentiation to myofibroblasts. The MUC1 cytoplasmic tail (CT) has also been implicated in intracellular signalling in fibrosis (33). The biology of MUC1 is complex, however, early trials in cancer, where MUC1 is a well-studied tumour-associated antigen, suggest that monoclonal antibodies or inhibitors of MUC1 might represent a new therapeutic avenue for IPF (34). Although the highest colocalisation probability implicated *MUC1*, the evidence was from blood only, and we cannot not rule out the possibility that another nearby gene might be the true causal gene for this locus. Of particular note, thrombospondin-3 (*THBS3*) has a known role in cell-to-cell and cell-to-matrix interactions and has been implicated in cardiac fibrosis (35) and skin healing (36).

The signal on chromosome 12, overlapping the 3’-UTR of *SNRPF*, provided the strongest colocalisation with gene expression evidence in this study for *NTN4* which encodes the secreted protein Netrin-4, originally identified as a guide of axon migration but more recently established to have an important role in kidney and vascular development, and has also been implicated in lung morphogenesis (37). *NTN4* is relatively highly expressed in aberrant basaloid cells (IPFCellAtlas.com; (38)), a recently described patient-specific cell population (39)(40)(41). *NTN4* has also been implicated by GWAS of diffusing capacity for carbon monoxide (DLCO) in a population enriched for COPD patients (42). The role of Netrin-4 in angiogenesis (43)(44)(45) and the renewed interest in the relevance of endothelial cell and vascular abnormalities in IPF (46) make this an intriguing signal for further investigation. Previous genetic association studies of other respiratory traits have also implicated Small Nuclear Ribonucleoprotein Polypeptide F (*SNRPF*) (47)(48) and centrosome gene, *CCDC38* (49) at this locus.

The most significant new signal (*p*=2.23×10^−11^) was located near to *LSM3*, encoding ‘LSM3 Homolog, U6 Small Nuclear RNA And MRNA Degradation Associated’, an sm-like protein with roles in RNA metabolism. The variant also mapped to *SLC6A6*, a taurine and beta-alanine transporter gene, implicated in Hypotaurinemic Retinal Degeneration and Cardiomyopathy (HTRDC) (50) and kidney fibrosis in diabetic knockout mice (51). Taurine may have a role in lung homeostasis and protection against oxidative stress (52)(53), a known driver of IPF pathogenesis.

All of the studies included in the genome-wide discovery stage of this analysis comprised cases that have been defined according to clinical criteria. Previous studies have noted an attenuation of effect sizes for IPF risk associated variants in studies where IPF case status has been defined using routine EHR (7)(54). Despite a smaller discovery sample size than previous studies that included IPF cases defined by EHR, we identified new signals that were replicated in an independent biobank EHR-based dataset (7). The use of the TOPMed Imputation reference panel enabled us to analyse over 25 million variants with a minor allele count greater than 3 and imputation quality greater than 0.5. This represented a more than 3-fold increase in variants analysed over our previous clinically-curated IPF GWAS (5). We included a larger number of variants than have been typically tested in GWAS; if we were to apply a more stringent genome-wide significance threshold of *p*<5×10^−9^ (for example, as applied in other recent large GWAS (55), two of our three new signals would have met this p-value threshold in our discovery GWAS and the third would also have exceeded this threshold had we applied a two-stage design with meta-analysis of discovery GWAS and replication data. Nevertheless, the consistency of the signals across the participating studies (including replication) provides reassurance on the robust nature of the signals.

A key challenge in translating genetic association signals to mechanistic insight is robust variant-to-gene mapping to understand the functional, and ultimately clinical, consequence of the genetic perturbation (56). We applied several *in silico* approaches to map the associated signals to genes. However, none of our signals included either a gene coding region or splice elements or had a high probability of sharing a causal variant with an eQTL for nearby genes. Additional functional *in silico* approaches, as well as *in vitro* and *in vivo* studies are needed to map these new signals to genes, predict their function, and investigate their biological function and relevance to IPF.

Our study only included individuals of European ancestry which means that we cannot assess the generalisability of these association signals to other populations. We were unable to validate several previously reported signals that had been reported in multi-ancestry studies. There is a dearth of studies of clinically-curated IPF case-control datasets; these prior multi-ancestry studies were predominantly derived from large population biobanks. Future efforts should be focused on increasing availability of carefully phenotyped IPF datasets from non-European ancestry populations.

In conclusion, we have identified new signals of association with IPF risk that may provide further mechanistic insight into the pathobiology of IPF and support identification of potential new drug targets.

## ETHICS

This research was conducted with appropriate ethics approval. The PROFILE study (which provided samples for the UK and UUS studies) had institutional ethics approval at the University of Nottingham (NCT01134822 - ethics reference 10/H0402/2) and Royal Brompton and Harefield NHS Foundation Trust (NCT01110694 - ethics reference 10/H0720/12). UK samples were recruited across multiple sites with individual ethics approval (University of Edinburgh Research Ethics Committee [The Edinburgh Lung Fibrosis Molecular Endotyping (ELFMEN) Study NCT04016181] 17/ES/0075, NRES Committee South West - Southmead, Yorkshire and Humber Research Ethics Committee 08/H1304/54 and Nottingham Research Ethics Committee 09/H0403/59). Spanish samples were recruited under ethics approval by ethics committee from the Hospital Universitario N.S. de Candelaria (reference of the approval: PI-19/12). The UUS study also included individuals from clinical trials with ethics approval (ACE [NCT00957242] and PANTHER [NCT00650091]). For the UCSF cohort, sample and data collection were approved by the University of California San Francisco Committee on Human Research and all patients provided written informed consent. For the Vanderbilt cohort, the Institutional Review Boards from Vanderbilt University approved the study and all participants provided written informed consent before enrolment. For individuals recruited at the University of Chicago, consenting patients with IPF who were prospectively enrolled in the institutional review board-approved ILD registry (IRB#14163A) were included. Individuals recruited at the University of Pittsburgh Medical Centre had ethics approval from the University of Pittsburgh Human Research Protection Office (referenceSTUDY20030223: Genetic Polymorphisms in IPF). Individuals from the COMET (NCT01071707) and Lung Tissue Research Consortium (NCT02988388) studies were also included in the Chicago study. All subjects in the Colorado study gave written informed consent as part of IRB-approved protocols for their recruitment at each site and the GWAS study was approved by the National Jewish Health IRB and Colorado Combined Institutional Review Boards (COMIRB). Subjects in the Genentech study provided written informed consent for whole-genome sequencing of their DNA. Ethical approval was provided as per the original clinical trials (INSPIRE [NCT00075998], RIFF [NCT01872689], CAPACITY [NCT00287729 and NCT00287716] and ASCEND [NCT01366209]). Individuals in the CleanUP-UCD study included individuals from clinical trials with ethics approval (NCT02759120). These samples were genotyped under University of Virginia ethics approval (IRB 20845). IPFJES involved human participants and was approved by East Midlands - Nottingham 1 Research Ethics Committee REC reference: 17/EM/0021IRAS project ID: 203355. Participants gave informed consent to participate in the study before taking part.

## DATA AVAILABILITY

Summary statistics (i.e., effect size estimates, standard errors, *p* values and basic variant information) for all variants included in the genome-wide meta-analysis can be accessed via https://github.com/genomicsITER/PFgenetics.

## AUTHORS CONTRIBUTION

LVW and RJA designed and supervised the study. DPC, TH-B, LD, BG-G, and OCL performed the analyses. AA, HLB, CleanUP-IPF Investigators of the Pulmonary Trials Cooperative, WAF, TEF, BG, IPH, SPH, MRH, NH, SRJ, NK, JML-S, S-FM, RJM, MIM, ADS, TMM, ABM, PLM, MM-M, VN, MN, JMO, HP, GS, IS, XRS, IDS, MES, MDT, MKBW, MCZ, YZ, FM, BLY, CJR, DAS, CF, IN, RGJ, and LVW participated in data collection. DPC, TH-B, and LVW wrote the first draft of the manuscript. All authors revised and approved the final version of the manuscript.

