## Supplementary material for "Genome-wide association study of Idiopathic Pulmonary Fibrosis susceptibility using clinically-curated European-ancestry datasets"

**Index**

### **SUPPLEMENTAL METHODS**

#### **Study samples**

We conducted an analysis from seven independent case-control studies of idiopathic pulmonary fibrosis (IPF) previously documented. For all seven studies, IPF cases were diagnosed based on the guidelines established by the American Thoracic Society and European Respiratory Society (1)(2)(3). The seven datasets are composed by Colorado (4), Chicago (5), UK (6), IPF Job Exposures Study (IPF-JES) (7), Genentech (8) (9), United States, United Kingdom, and Spain (UUS) (10), and Study of Clinical Efficacy of Antimicrobial Therapy Strategy Using Pragmatic Design in Idiopathic Pulmonary Fibrosis - University of California, Davis (CleanUP-UCD) (11).

The Colorado study (4), a cohort of 1,515 patients with fibrotic idiopathic interstitial pneumonias (fIIP), was sourced from various US cohorts (including National Jewish Health IIP population, InterMune IPF trials, UCSF, Vanderbilt University IIP population, and the National Heart Lung and Blood Institute Lung Tissue Research Consortium). These cases were matched with 2,455 population controls chosen for their genetic similarity. Genotyping for all individuals was conducted using the Illumina Human 660W Quad BeadChip array.

The US study (previously referred as "Chicago" study) (5) encompassed 541 IPF cases from the University of Chicago, University of Pittsburgh, and the COMET study. It also included 542 population controls from the database of genotypes and phenotypes (dbGaP) and the University of Pittsburgh. Genotyping for both cases and controls was carried out using the Affymetrix Genome-Wide Human SNP 6.0 array.

The UK study (6) gathered a total of 612 IPF cases from 9 different centres across the UK, and 3,366 matching controls selected from the UK Biobank. Cases were genotyped with the Affymetrix UK BILEVE array, while controls were genotyped with the UK Biobank array.

The Idiopathic Pulmonary Fibrosis Job Exposures Study (IPF-JES) (7) included 416 men from England, Scotland, or Wales diagnosed with IPF, along with 2,465 matched male controls recruited to the IPF-JES study or selected from UK Biobank. Genotyping was carried out using the Affymetrix UK Biobank array.

The Genentech study (8) (9) comprised 813 cases derived from three IPF clinical trials (ASCEND, CAPACITY, and RIFF) and 3,949 controls from four non-IPF clinical trials including age-related macular degeneration, diabetic macular oedema, multiple sclerosis, asthma, and inflammatory bowel disease. Genotyping was accomplished through whole-genome sequencing using the HiSeq X Ten platform (Illumina) with an average read depth of 30X. Individuals were filtered with ADMIXTURE v1.23 to keep only the European ancestry individuals, and also by missingness, relatedness and heterozygosity. Variants were filtered according to missingness >0.1, Hardy-Weinberg Equilibrium (HWE) (*p*=5x10^-8^), case-control missingness and allele balance.

The UUS study (United States, United Kingdom, and Spain) (10) included 793 IPF cases from 7 study cohorts (ACE, PANTHER, UCD, Chicago, UCSF, PROFILE, and Spain), along with 9,999 population controls selected from the UK Biobank to match ancestry, sex, and smoking distribution. Genotyping for cases was done using the Affymetrix UK Biobank and Spanish Biobank arrays, while controls were genotyped with the UK Biobank array.

The CleanUP-UCD study (11), encompassed a total of 469 IPF cases drawn from a randomized clinical trial conducted across 35 locations in the United States, and 32 participants were recruited at University of California Davis (UCD). Additionally, 2,455 population controls were selected from the UK Biobank. Genotyping was performed on both cases and controls using the Affymetrix UK Biobank array.

Six case-control studies (Colorado, IPF-JES, UK, US, UUS, and CleanUP-UCD) were imputed using the TOPMed WGS reference panel (GRCh38) via the TOPMed Imputation Server, ensuring that only SNPs available in both (cases and controls) were included.

For the X chromosome, we performed the analysis of four independent case-control studies, including Colorado, UK, UUS and CleanUP-UCD. Genotyping and quality control were performed as previously described (following previously described methods: HWE, CR95% and MAF) and imputed using TOPMed. We performed association analysis adjusted for 10 principal components using PLINK v2 (12) with the option --xchr-model 2, to consider the males and female dosages on a 0-2 scale. Variants with a poor imputation quality (r2<0.5) and a low minor allele frequency (MAF<0.01) were removed. We meta-analysed results using inverse-variance weighted fixed effect meta-analysis using METAL (13) and excluded SNPs that were not measured in at least two studies. We estimated the genomic inflation factor using the classical lambda and corrected by this value.

#### **Statistical analysis**

Bayesian fine-mapping

Fine mapping was used to generate a set of genetic variants with a 95% probability of containing the causal variant associated with a given genetic signal (95% credible set). This process was performed using R version 4.3.1. The fine-mapping strategy used, using the Wakefield approximate Bayes factor (14), assumed the existence of a single measured causal variant. The variants from the credible set were annotated using ANNOVAR v07.06.20 (15).

Colocalisation

For those variants showing associations with gene expression levels, colocalisation analyses were performed in lung, cultured fibroblasts, and whole blood tissues (Genotype-Tissue Expression Project: GTEx) corresponding to the relevant genes. These analyses were performed using the coloc (16) package in R version 4.3.1.

Colocalisation analyses were used to determine whether the same causal variant was associated with both IPF susceptibility and gene expression in GTEx. The posterior probability was estimated using the approximate Bayes factor:

- H0: neither IPF susceptibility nor gene expression have a genetic association in that region
- H1: only IPF susceptibility has a genetic association in that region
- H2: only gene expression has a genetic association in that region
- H3: both IPF susceptibility and gene expression are associated, but with different causal variants
- H4: both IPF susceptibility and gene expression are associated and share a single causal variant

If the posterior probability in favour of the alternative hypothesis, indicating the presence of a common single causal variant for both IPF susceptibility and gene expression (H4), exceeded 70%, we inferred that the IPF and gene expression signals showed colocalisation.

#### **Functional annotation**

We conducted a thorough functional impact assessment using empirical data from several integrated software tools and datasets. We used RegulomeDB (17) and The Open Targets Post-GWAS portal (18) to assess the regulatory potential and prioritise the functional roles of each SNP and its associated gene. In addition, we used Open Targets (<https://www.opentargets.org/>) (18) to access cross-referencing of genetic variants with a wide range of phenotypes. Variants were annotated using various databases for variant type, protein function and pathogenic potential using Ensembl VEP v105.

Active chromatin marks

We also functionally annotated the highlighted regions based on active chromatin marks, including DNase I hypersensitivity hotspots (DHS), open chromatin peaks, transcription factor footprints, enhancer RNAs (eRNAs), Assay for Transposase-Accessible Chromatin using sequencing (ATACseq), ChIP sequencing (CHIPseq), and epigenetic modification of the DNA packaging protein histones (H3K27ac) (19).

Mouse-knockout orthologs with a respiratory phenotype

We selected human orthologs of mouse knockout genes with respiratory phenotypes listed in the International Mouse Phenotyping Consortium (<https://www.mousephenotype.org/>), within ±500 kb of a IPF sentinel variant.

### **SUPPLEMENTAL TABLES**

#### **Table S1.** Sample size of the study.

|  | **Cases** | **Controls** | **Autosomal variants analysed** | **X chromosome variants analysed** |
| --- | --- | --- | --- | --- |
| Colorado | 1,515 | 4,683 | 24,275,956 | 238,835 |
| US | 541 | 542 | 13,539,842 | --- |
| UK | 612 | 3,365 | 19,550,111 | 262,880 |
| IPF-JES | 416 | 2,465 | 18,681,239 | --- |
| Genentech | 813 | 3,949 | 14,955,256 | --- |
| UUS | 793 | 9,999 | 33,838,128 | 255,656 |
| CleanUP-UCD | 469 | 2,455 | 19,500,953 | 262,831 |
| Total | 5,159 | 27,458 | 25,290,839 | 261,736 |
| The number of individuals and variants shown is the result after quality controls. | | | | |

#### **Table S2.** Bayesian fine mapping results from new signals sorted by posterior inclusion probability.

| **CHR** | **BP** | **rsID** | **Location** | **Nearest genes** | **A1*/A2** | **A1 Freq.** | **OR (CI95%)** | **P** | **PIP (95%)** |
| --- | --- | --- | --- | --- | --- | --- | --- | --- | --- |
| 1 | 155179278 | rs9426886 | intronic | *TRIM46* | A/T | 0.568 | 0.861 (0.820-0.904) | 1.53x10^-9^ | 0.208 |
| 1 | 155179017 | rs11264341 | intronic | *TRIM46* | T/C | 0.432 | 1.161 (1.106-1.219) | 1.70x10^-9^ | 0.189 |
| 1 | 155204315 | rs2075571 | intronic | *THBS3* | T/C | 0.428 | 1.161 (1.106-1.219) | 1.83x10^-9^ | 0.176 |
| 1 | 155183255 | rs4971100 | intronic | *TRIM46* | A/G | 0.430 | 1.160 (1.105-1.218) | 2.17x10^-9^ | 0.149 |
| 1 | 155185239 | rs2070803 | downstream | *MUC1\TRIM46* | A/G | 0.567 | 0.864 (0.823-0.907) | 3.74x10^-9^ | 0.089 |
| 1 | 155199564 | rs423144 | intronic | *THBS3* | T/G | 0.429 | 1.157 (1.103-1.215) | 3.74x10^-9^ | 0.089 |
| 1 | 155185159 | rs4971101 | downstream | *MUC1\TRIM46* | A/G | 0.432 | 1.156 (1.101-1.214) | 4.77x10^-9^ | 0.070 |
| 3 | 14270502 | rs112271207 | intergenic | *LSM3\LINC01267* | T/C | 0.060 | 1.356 (1.231-1.494) | 2.23x10^-11^ | 0.691 |
| 3 | 14269725 | rs114455129 | intergenic | *LSM3\LINC01267* | T/C | 0.051 | 0.743 (0.676-0.817) | 1.36x10^-10^ | 0.118 |
| 3 | 14301732 | rs116678873 | intergenic | *LSM3\LINC01267* | A/G | 0.058 | 0.744 (0.677-0.818) | 6.81x10^-10^ | 0.029 |
| 3 | 14298964 | rs111405596 | intergenic | *LSM3\LINC01267* | T/C | 0.939 | 1.335 (1.216-1.465) | 8.45x10^-10^ | 0.024 |
| 3 | 14303378 | rs112304124 | intergenic | *LSM3\LINC01267* | A/G | 0.939 | 1.342 (1.221-1.476) | 1.10x10^-9^ | 0.019 |
| 3 | 14308107 | rs112255370 | intergenic | *LSM3\LINC01267* | T/C | 0.064 | 1.334 (1.215-1.464) | 1.14x10^-9^ | 0.019 |
| 3 | 14304955 | rs113247615 | intergenic | *LSM3\LINC01267* | A/C | 0.061 | 0.745 (0.677-0.819) | 1.17x10^-9^ | 0.018 |
| 3 | 14308058 | rs111345960 | intergenic | *LSM3\LINC01267* | T/C | 0.064 | 0.811 (0.754-0.872) | 1.24x10^-9^ | 0.017 |
| 3 | 14305318 | rs111275427 | intergenic | *LSM3\LINC01267* | T/C | 0.939 | 1.233 (1.147-1.326) | 1.24x10^-9^ | 0.017 |
| 12 | 95866696 | rs7957346 | 3’UTR | *SNRPF* | A/C | 0.582 | 0.811 (0.753-0.872) | 1.56x10^-8^ | 0.376 |
| 12 | 95869659 | rs71443541 | intronic | *CCDC38* | A/G | 0.583 | 0.870 (0.829-0.914) | 2.35x10^-8^ | 0.254 |
| 12 | 95884368 | rs4762250 | intronic | *CCDC38* | A/G | 0.435 | 1.145 (1.091-1.203) | 3.70x10^-8^ | 0.164 |
| 12 | 95860750 | rs6538677 | intronic | *SNRPF* | A/G | 0.398 | 1.145 (1.091-1.203) | 5.15x10^-8^ | 0.121 |
| 12 | 95837440 | rs76152842 | ncRNA_intronic | *LOC105369921* | T/C | 0.608 | 0.876 (0.834-0.920) | 1.06x10^-7^ | 0.063 |
| *A1: effect allele; OR: Odds Ratio; PIP: posterior inclusion probability; Coordinates in GRCh38, Freq.: frequency; CI: Confidence Interval. | | | | | | | | | |

#### **Table S3.** Functional assessment of sentinels and variants with the posterior inclusion probability (PIP)>0.1 in the credible set.

| **Region**  **(PIP>0.1)** | **Nereast genes** | **rsID** | **Location** | **V2G** | **CADD** | **LoFtool** | **Colocalisation eQTL (GTEx)**  **[highest PP-H4]** | **RegulomeDB / score** | **PheWAS (p<5X10^-8^)**  **[Or most significant]** | **Mouse KO** | **Active mark annotation** | **eRNA or footprints** |
| --- | --- | --- | --- | --- | --- | --- | --- | --- | --- | --- | --- | --- |
| 1q22  (0.208) | *TRIM46* | rs9426886 | intronic | *THBS3*  (sQTL; eQTL) | 4.64 | 0.022 | Coloc=61.6% in whole blood (*MUC1*); | eQTL/caQTL + TF binding / chromatin accessibility peak (0.66703) | Hematocrit measurement (*p*=4.7x10^-41^) | None | DHS; ATACseq; H3K27ac | No |
| 1q22  (0.189) | *TRIM46* | rs11264341 | intronic | *ADAM15*  (sQTL; eQTL; DHS-promoter correlation) | 3.04 | 0.022 |  | eQTL/caQTL + TF binding / chromatin accessibility peak (0.55436) | Urate levels measurement (*p*=1.5x10^-101^) | None | DHS; ATACseq; H3K27ac | No |
| 1q22  (0.176) | *TRIM46* | rs4971100 | intronic | *THBS3*  (sQTL; eQTL) | 3.33 | None |  | eQTL/caQTL + TF binding / chromatin accessibility peak (0.55436) | Hematocrit measurement (*p*=1.2x10^-40^) | None | H3K27ac | No |
| 1q22  (0.149) | *THBS3* | rs2075571 | intronic | *THBS3*  (sQTL; eQTL) | 3.01 | 0.337 |  | eQTL/caQTL + TF binding / chromatin accessibility peak (0.66703) | Hematocrit measurement (*p*=6.8x10^-41^) | None | DHS; H3K27ac | No |
| 3p25.1  (0.691) | *LSM3*\ *LINC01267* | rs112271207 | intergenic | *SLC6A6*  (PCHi-C) | 7.51 | None | No significant eQTLs | TF binding or chromatin accessibility peak  (0.13454) | Lung function (FEV1/FVC) measurement (*p*=6.1x10^-8^) | None | ATACseq; H3K27ac | No |
| 3p25.1  (0.118) | *LSM3*\ *LINC01267* | rs114455129 | intergenic | *SLC6A6*  (PCHi-C) | 0.195 | None |  | TF binding + chromatin accessibility peak  (0.609069) | Lung function (FEV1/FVC) measurement (*p*=1.5x10^-5^) | None | ATACseq; H3K27ac | No |
| 12q23.1  (0.376) | *SNRPF* | rs7957346 | 3’UTR | *NTN4*  (sQTL) | 2.9 | None | Coloc=64.9% in lung (*NTN4*) | eQTL/caQTL + TF binding / chromatin accessibility peak (0.55324) | Lung function (FEV1/FVC) measurement (*p*=7.8x10^-45^) | None | ATACseq; H3K27ac | No |
| 12q23.1  (0.254) | *SNRPF* | rs6538677 | intronic | *NTN4*  (sQTL) | 0.03 | None |  | eQTL/caQTL + TF binding / chromatin accessibility peak (0.55324) | Lung function (FEV1/FVC) measurement (*p*=2.1x10^-38^) | None | ATACseq; H3K27ac | No |
| 12q23.1  (0.164) | *CCDC38* | rs10735335 | intronic | *NTN4*  (sQTL) | 1.66 | 0.974 |  | eQTL/caQTL + TF binding / chromatin accessibility peak (0.68) | Lung function (FEV1/FVC) measurement (*p*=2.2x10^-35^) | None | DHS; ATACseq; H3K27ac | No |
| 12q23.1  (0.121) | *CCDC38* | rs4762250 | intronic | *NTN4*  (sQTL) | 11.4 | 0.974 |  | eQTL/caQTL + TF binding / chromatin accessibility peak (0.55324) | Lung function (FEV1/FVC) measurement (*p*=6.9x10^-33^) | None | H3K27ac | No |
| CADD: Combined annotation dependent depletion; DHS: DNase I sensitivity quantitative trait loci; eQTL: Expression quantitative trait loci; eRNA: enhancer RNAO: Knockout; LoFtool: Gene intolerance score based on loss-of-function variants, LoFtool score < 0.1 low tolerance to loss function; PheWAS: Related traits from Open Target Genetics; PP-H4; posterior probability; RegulomeDB model score is ranging from 0 to 1, with 1 being most likely to be a regulatory variant; sQTL: Splicing quantitative trait loci; V2G: Automated variant to gene assignment from Open Targets Genetics. | | | | | | | | | | | | |

#### **Table S4.** Association results for sentinels previously reported genome-wide signals.

| **CHR** | **BP** | **rsID** | **Location** | **Nearest gene** | **A1*/A2** | **A1 Freq.** | **OR (CI95%)** | **P** | **Firstly reported^+^** |
| --- | --- | --- | --- | --- | --- | --- | --- | --- | --- |
| 1 | 9061448 | rs2896012 | intronic | *SLC2A5* | A/T | 0.611 | 0.88 (0.84-0.92) | **3.21x10^-7^** | Partanen (2022) [PMID: 36777997] |
| 1 | 77998184 | rs4130548 | intronic | *DNAJB4* | T/C | 0.634 | 0.95 (0.91-1.00) | 0.063 | Partanen (2022) [PMID: 36777997] |
| 1 | 150579566 | rs16837903 | 5’ UTR | *MCL1* | A/G | 0.144 | 0.81 (0.75-0.87) | **1.31x10^-8^** | Peljto (2023)  [PMID: 36602845] |
| 1 | 214482547 | rs12096551 | intronic | *PTPN14* | T/C | 0.801 | 1.22 (1.15-1.30) | 6.96x10^-10^ | Allen (2022)  [PMID: 35688625] |
| 3 | 44804157 | rs74341405 | intronic | *KIF15* | T/C | 0.952 | 0.63 (0.57-0.70) | 7.10x10^-19^ | Allen (2020)  [PMID: 31710517] |
| 3 | 169769713 | rs9811216 | upstream | *TERC* | T/C | 0.729 | 0.77 (0.74-0.82) | **3.27x10^-21^** | Fingerlin (2013) [PMID: 23583980] |
| 4 | 88892133 | rs6815970 | intronic | *FAM13A* | T/C | 0.228 | 1.31 (1.24-1.38) | **1.88x10^-21^** | Fingerlin (2013) [PMID: 23583980] |
| 4 | 159892716 | rs76537958 | intergenic | *FSTL5* | A/T | 0.966 | 0.84 (0.74-0.96) | 9.47x10^-3^ | Partanen (2022) [PMID: 36777997] |
| 5 | 1286401 | rs2736100 | intronic | *TERT* | A/C | 0.510 | 1.37 (1.30-1.44) | **7.86x10^-36^** | Mushiroda (2008) [PMID: 18835860] |
| 5 | 169588475 | rs116483731 | exonic | *SPDL1* | A/G | 0.010 | 2.79 (2.29-3.39) | 1.14x10^-24^ | Dhindsa (2021) [PMID: 33758299] |
| 6 | 7562999 | rs2076295 | intronic | *DSP* | T/G | 0.530 | 0.68 (0.65-0.72) | 4.81x10^-53^ | Fingerlin (2013) [PMID: 23583980] |
| 6 | 35549503 | rs9348978 | intergenic | *FKBP5* | A/G | 0.518 | 0.87 (0.83-0.91) | **1.27x10^-8^** | Partanen (2022) [PMID: 36777997] |
| 6 | 43386693 | rs1214761 | intergenic | 6p21.2 | A/G | 0.323 | 0.85 (0.81-0.90) | **1.32x10^-9^** | Allen (2022)  [PMID: 35688625] |
| 7 | 1936920 | rs2280550 | intronic | *MAD1L1* | A/G | 0.377 | 0.79 (0.75-0.83) | **7.15x10^-20^** | Allen (2020)  [PMID: 31710517] |
| 7 | 100020229 | rs35228488 | intronic | *ZKSCAN1* | C/G | 0.384 | 1.28 (1.22-1.35) | **1.50x10^-23^** | Allen (2022)  [PMID: 35688625] |
| 7 | 129095384 | rs34288126 | intergenic | *TPI1P2* | A/G | 0.129 | 1.12 (1.04-1.20) | 1.63x10^-3^ | Partanen (2022) [PMID: 36777997] |
| 8 | 119927966 | rs10808505 | intronic | *DEPTOR* | T/G | 0.572 | 1.20 (1.14-1.26) | **4.78x10^-13^** | Allen (2020)  [PMID: 31710517] |
| 9 | 106721382 | rs1388233 | intergenic | 9q31.2 | A/G | 0.519 | 1.17 (1,12-1.23) | **1.26x10^-10^** | Allen (2022)  [PMID: 35688625] |
| 10 | 103882177 | 10:103882177 | 3’UTR | *OBFC1* | G/^~^ | 0.493 | 0.85 (0.81-0.90) | **2.14x10^-10^** | Allen (2022)  [PMID: 35688625] |
| 10 | 109470103 | rs79684490 | intergenic | *10q25.1* | A/G | 0.047 | 1.29 (1.16-1.43) | **3.10x10^-6^** | Allen (2022)  [PMID: 35688625] |
| 11 | 1219991 | rs35705950 | intergenic | *MUC5B* | T/G | 0.150 | 5.22 (4.89-5.58) | **0^#^** | Seibold (2011)  [PMID: 21506741] |
| 13 | 112886111 | rs3742238 | 3’ UTR | *ATP11A* | T/C | 0.206 | 0.78 (0.74-0.83) | 3.22x10^-14^ | Fingerlin (2013) [PMID: 23583980] |
| 15 | 40424054 | rs2304645 | intronic | *IVD* | C/G | 0.526 | 1.27 (1.21-1.33) | **1.10x10^-21^** | Fingerlin (2013) [PMID: 23583980] |
| 15 | 40621642 | rs2412541 | exonic | *KNL1* | T/G | 0.840 | 0.79 (0.74-0.84) | **1.66x10^-13^** | Allen (2022)  [PMID: 35688625] |
| 15 | 85744679 | rs11073517 | 3’UTR | *AKAP13* | T/C | 0.324 | 1.18 (1.12-1.24) | **2.42x10^-10^** | Allen (2017)  [PMID: 29066090] |
| 16 | 34035 | rs367849850 | intergenic | *IL9RP3* | G/^$^ | 0.042 | 2.28 (2.03-2.55) | 7.82x10^-47^ | Donoghue (2023) [PMID: 36603154] |
| 16 | 67895674 | rs539683219 | intronic | *PSKH1* | TG/T | NA | NA | NA | Partanen (2022) [PMID: 36777997] |
| 17 | 45949182 | rs242561 | intronic | *KANSL1* (17q21.31) | T/C | 0.221 | 0.68 (0.64-0.73) | **1.23x10^-31^** | Fingerlin (2013) [PMID: 23583980] |
| 19 | 4717660 | rs12610495 | intronic | *DPP9* | A/G | 0.694 | 0.77 (0.73-0.81) | **1.00x10^-23^** | Fingerlin (2013) [PMID: 23583980] |
| 19 | 5840608 | rs708686 | intergenic | *FUT6* | T/C | 0.281 | 1.18 (1.12-1.25) | 1.05x10^-9^ | Partanen (2022) [PMID: 36777997] |
| 20 | 63652817 | rs112087793 | intronic | *STMN3* | T/C | 0.083 | 0.78 (0.71-0.86) | **2.40x10^-7^** | Allen (2022)  [PMID: 35688625] |
| 20 | 63694480 | rs115610405 | exonic | *RTEL1* | A/C | 0.022 | 1.64 (1.41-1.92) | 2.01x10^-10^ | Allen (2022)  [PMID: 35688625] |
| *A1: effect allele; OR: Odds Ratio. ^#^p<5.00x10^-121^  ^~^GCAAGTGA  ^$^GGGGAGCCTGGAAGCACAC Coordinates in GRCh38; Freq.: Frequency; CI: Confidence Interval.  ^+^Mushiroda et al 2008 (20); Seibold et al 2011 (21); Fingerlin et al 2013 (4); Allen et al 2017 (6); Allen et al 2020 (10); Dhindsa et al 2021 (22); Allen et al 2022 (23); Partanen et al 2022 (24); Donoghue et al 2023 (8); Peljto et al 2023 (9). | | | | | | | | | |

#### **Table S5.** Colocalisation analysis between the IPF risk signals and the gene expression eQTL signals in lung, fibroblasts and blood.

| **Region** | **Gencode ID**  **(Gene Symbol)** | **SNP-effect allele (IPF risk allele)** | **P** | **NES** | **Tissue** | **PP.H0.abf** | **PP.H1.abf** | **PP.H2.abf** | **PP.H3.abf** | **PP.H4.abf** |
| --- | --- | --- | --- | --- | --- | --- | --- | --- | --- | --- |
| 1q22 | ENSG00000160766.14 (*GBAP1*) | rs9426886-T (T) | 5.1x10^-25^ | -0.43 | Cells cultured fibroblasts | 3.62x10^-39^ | 1.40x10^-35^ | 2.56x10^-4^ | 0.988 | 0.012 |
|  | ENSG00000160766.14 (*GBAP1*) | rs9426886-T (T) | 2.7x10^-20^ | -0.28 | Lung | 2.45x10^-34^ | 9.44x10^-31^ | 2.56x10^-4^ | 0.987 | 0.012 |
|  | ENSG00000169231.13 (*THBS3*) | rs9426886-T (T) | 4.4x10^-20^ | 0.23 | Whole blood | 2.76x10^-64^ | 1.06x10^-60^ | 2.58x10^-4^ | 0.994 | 6.02x10^-3^ |
|  | ENSG00000169231.13 (*THBS3*) | rs9426886-T (T) | 1x10^-9^ | 0.19 | Lung | 4.28x10^-21^ | 1.65x10^-17^ | 2.58x10^-4^ | 0.996 | 4.10x10^-3^ |
|  | **ENSG00000185499.16 (*MUC1*)** | **rs9426886-T (T)** | **1.5x10^-9^** | **0.22** | **Whole blood** | **7.02x10^-10^** | **2.71x10^-6^** | **9.96x10^-5^** | **0.383** | **0.616** |
|  | ENSG00000236263.1 (*RP11-263K19.6*) | rs9426886-T (T) | 5.5x10^-9^ | 0.13 | Whole blood | 8.10x10^-12^ | 3.12x10^-8^ | 2.53x10^-4^ | 0.974 | 0.026 |
|  | ENSG00000160766.14 (*GBAP1*) | rs9426886-T (T) | 1.8x10^-8^ | -0.21 | Whole blood | 5.03x10^-17^ | 1.94x10^-13^ | 2.56x10^-4^ | 0.985 | 0.015 |
|  | ENSG00000169241.17 (*SLC50A1*) | rs9426886-T (T) | 3.7x10^-8^ | 0.13 | Cells cultured fibroblasts | 8.60x10^-15^ | 3.31x10^-11^ | 2.57x10^-4^ | 0.990 | 0.010 |
|  | ENSG00000231064.7 (*RP11-263K19.4*) | rs9426886-T (T) | 2.40x10^-7^ | 0.19 | Whole blood | 2.12x10^-15^ | 8.15x10^-12^ | 2.58x10^-4^ | 0.996 | 3.89x10^-3^ |
|  | ENSG00000143537.13 (*ADAM15*) | rs9426886-T (T) | 9.30x10^-7^ | -0.12 | Cells cultured fibroblasts | 5.32x10^-17^ | 2.05x10^-13^ | 1.44x10^-4^ | 0.553 | 0.447 |
|  | ENSG00000231064.7 (*RP11-263K19.4*) | rs9426886-T (T) | 2.00x10^-6^ | 0.14 | Lung | 2.40x10^-7^ | 9.24x10^-4^ | 1.41x10^-4^ | 0.544 | 0.455 |
|  | ENSG00000225855.6 (*RUSC1-AS1*) | rs9426886-T (T) | 2.80x10^-6^ | 0.11 | Cells cultured fibroblasts | 1.29x10^-15^ | 4.97x10^-12^ | 2.59x10^-4^ | 1.000 | 3.82x10^-5^ |
|  | ENSG00000169242.11 (*EFNA1*) | rs9426886-T (T) | 5.50x10^-5^ | -0.12 | Lung | 1.53x10^-7^ | 5.91x10^-4^ | 2.50x10^-4^ | 0.964 | 0.035 |
| 12q23.1 | ENSG00000258343.1 (*RP11-536G4.2*) | rs4762250-G (A) | 6.20x10^-19^ | -0.48 | Lung | 8.63x10^-73^ | 1.85x10^-70^ | 2.57x10^-3^ | 0.549 | 0.448 |
|  | ENSG00000139343.10 (*SNRPF*) | rs4762250-G (A) | 3.30x10^-7^ | -0.087 | Whole blood | 1.01x10^-6^ | 2.15x10^-4^ | 3.53x10^-3^ | 0.754 | 0.242 |
|  | ENSG00000139344.7 (*AMDHD1*) | rs4762250-G (A) | 2.10x10^-5^ | -0.18 | Lung | 1.69x10^-6^ | 3.61x10^-4^ | 3.21x10^-3^ | 0.686 | 0.310 |
|  | **ENSG00000074527.11 (*NTN4*)** | **rs6538677-G (A)** | **4.00x10^-5^** | **0.13** | **Lung** | **2.88x10^-4^** | **0.062** | **1.35x10^-3^** | **0.287** | **0.649** |
| NES: normalized effect size. | | | | | | | | | | |

### **SUPPLEMENTAL FIGURES**

**Figure S1. Quantile-quantile plot** of the observed (y-axis) versus expected -log10 p-values (x-axis) of the meta-analysis results (before genomic control correction): a) autosomal analysis (LDsc inflation factor λ=1.18); b) X chromosome (classic inflation factor λ=1.26).

a)

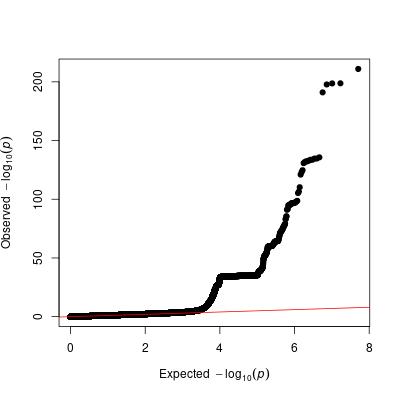

b)

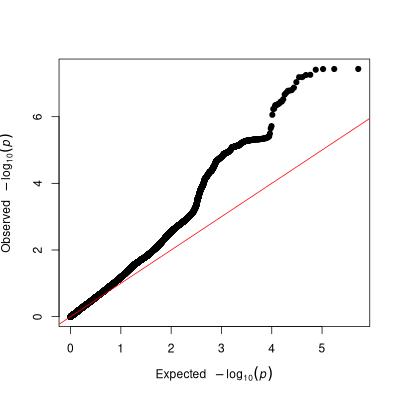

**Figure S2. Mirror plots** of the signals of interest with the highest posterior probability H4: a) 1q22 region, rs9426886 (intronic to *TRIM46*); and b) 12q23.1 region, rs7957346 (3’UTR to *SNRPF*). The green box represents the gene location.

a) 1q22 region, rs9426886 (intronic to *TRIM46*): *MUC1* in whole blood. IPF association results above and gene expression below.

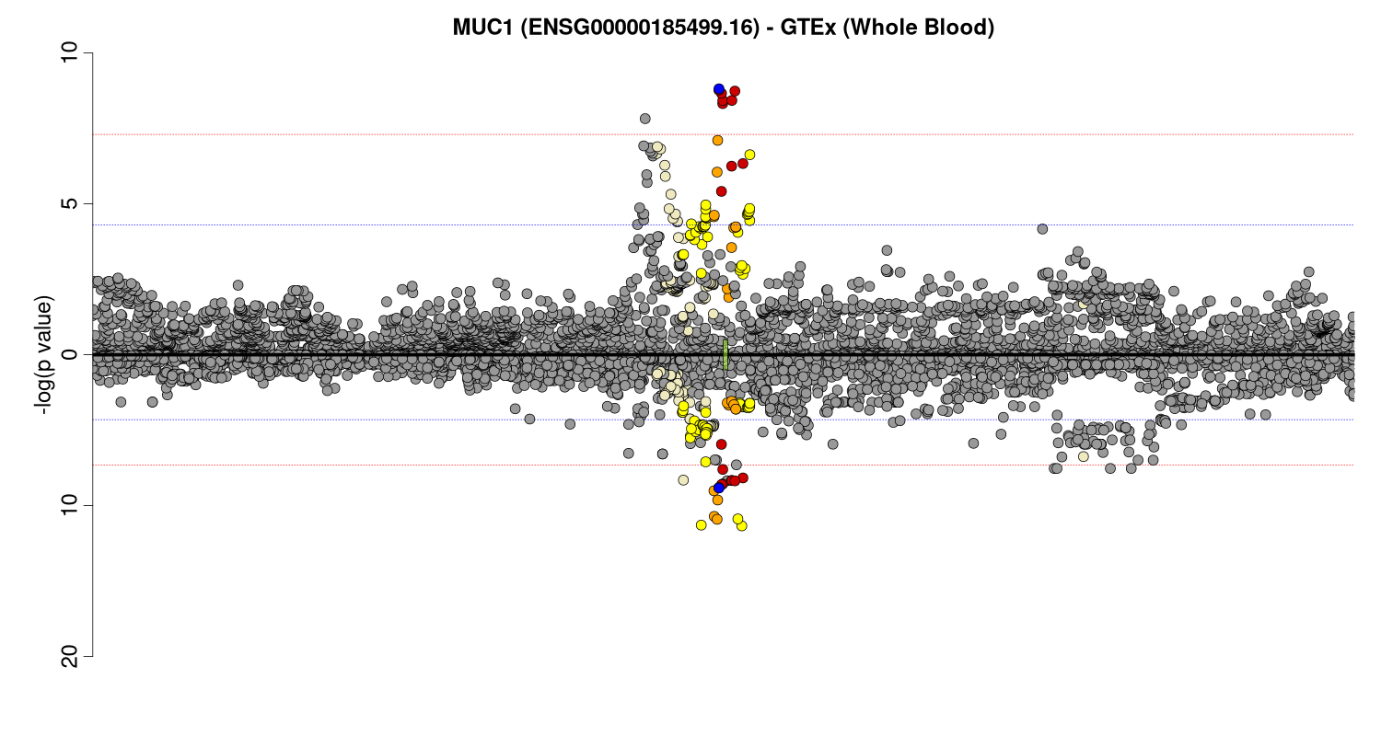

b) 12q23.1 region, rs7957346 (3’UTR to *SNRPF*): *NTN4* in lung. IPF association results above and gene expression below.

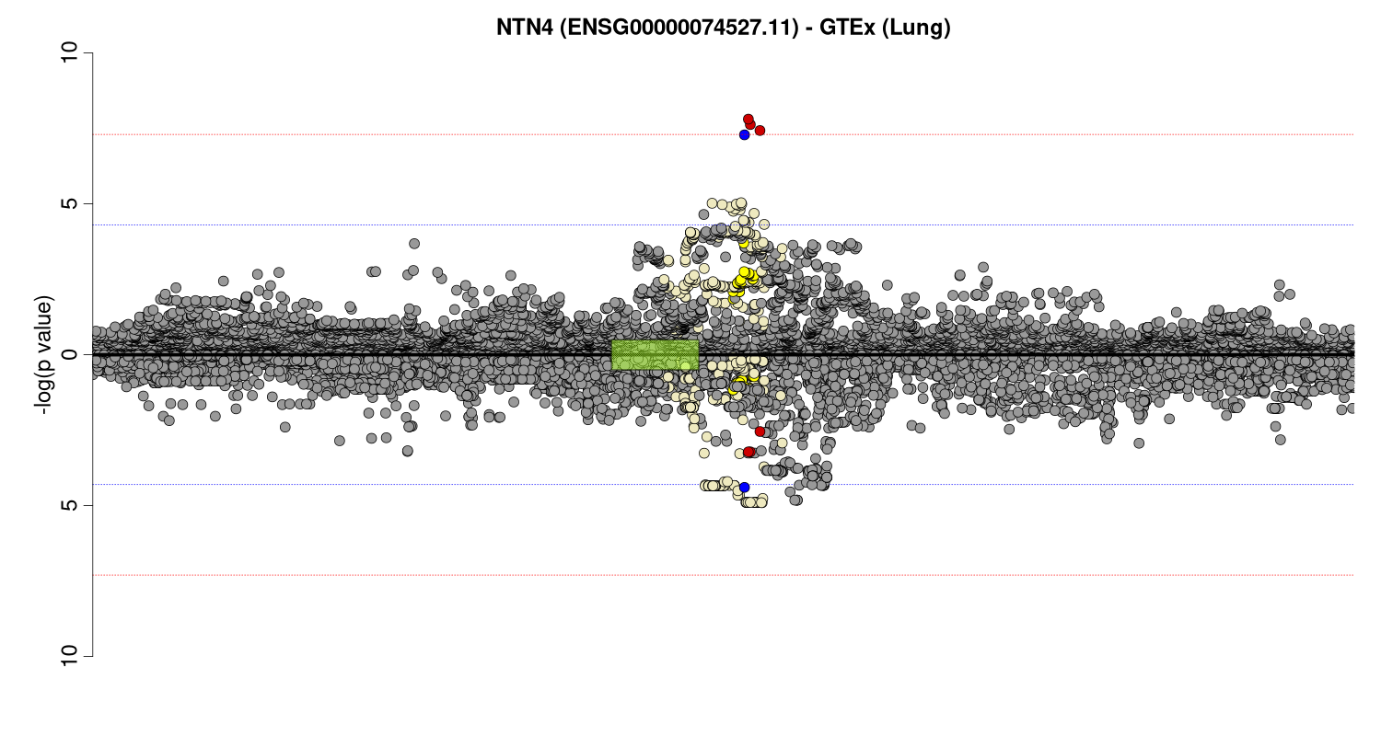
